# Deciphering the cell-intrinsic effect of DNMT3A clonal hematopoiesis in circulating monocytes: potential mechanistic insights into a protective role in Alzheimer’s disease

**DOI:** 10.1101/2023.09.29.23296147

**Authors:** Wesley T. Abplanalp, Stefanie Dimmeler, Andreas M. Zeiher

## Abstract

**Background:** Recent evidence showed an unforeseen connection between clonal hematopoiesis (CH) and protection against Alzheimer’s disease (AD) and found mutant bone marrow-derived cells infiltrate the brain and acquire a microglial-like phenotype, correlating with reduced neuritic plaques and neurofibrillary tangles in non-AD individuals. CH, characterized by somatic mutations in hematopoietic stem cells leading to clonal expansion of immune cells, has previously been associated with the exacerbation of various age-related diseases.

**Methods and Results:** As the mechanism behind this protective effect remains unclear, our study introduces a novel method, MutDetect-Seq, capable of discerning cell-intrinsic effects of CH mutations. Single-cell RNA sequencing profiles in patients with DNMT3A CH mutations were assessed, hypothesizing that mutant cells might augment phagocytosis, potentially mitigating neurodegenerative processes. Indeed, DNMT3A mutant monocytes exhibited a distinct pro-phagocytic gene signature as demonstrated via upregulation of prototypical phagocytotic genes like CALR, FCGR1A, CYBA, S100A8, S100A9 and FCER1G. Upregulation was validated in vitro with DNMT3A silenced THP1 macrophages. Furthermore, upregulated gene pathways in mutant monocytes associated with phagocytosis, suggesting bone marrow-derived monocytic cells with CH DNMT3A mutations could replenish the brain’s phagocytic capacity, counteracting age-related declines in microglial phagocytosis, and thereby causatively reducing neurodegenerative processes in AD.

**Conclusion:** This study reveals critical phenotypic disparities between DNMT3A CH driver harboring mutant and wild-type monocytes, shedding light on a potential mechanism underlying the unexpected protective role of CH in AD progression and offering insights into the effects of different CH mutations in disease contexts.

---

Recently, Jaiswal and colleagues^1^ reported in *Nature Medicine* a seminal and unexpected discovery: clonal hematopoiesis (CH) is associated with protection from incident Alzheimer’s disease (AD). In recent years, CH, defined as the occurrence of specific somatic mutations in hematopoietic stem cells leading to clonal expansion of circulating immune cells, was shown to associate with the promotion and worse outcomes of various of aging-associated diseases, including cardiovascular diseases, osteoporosis, chronic obstructive pulmonary disease, cancer and haematological malignancies.^2^ AD, a leading cause of morbidity and mortality among the elderly,^3^ appeared to be another addition to this list of ailments. Yet, contrary to expectations and quite surprisingly, Jaiswal and colleagues^1^ not only present genetic evidence of a negative association between AD risk and CH but, more importantly, demonstrate that mutant, bone marrow-derived cells infiltrate the brain and adopt a microglial-like phenotype. Intriguingly, the presence of these mutated cells in the brain parenchyma correlates with lower levels of neuritic plaques and neurofibrillary tangles in individuals without AD. Despite these suggestive findings, the authors acknowledge that their data do not offer any mechanistic insights into how CH mutant cells might impede neurodegenerative processes in the brain to protect against incident AD. To establish a potential causal role in AD protection, identifying biologically relevant differences between mutant and wild-type cells would be essential.

We have recently established a novel method to identify cell-intrinsic effects of CH (termed *MutDetect-Seq*), which for the first time enables the differentiation of mutant versus wild-type immune cells in patients harbouring CH mutations.^4^ Inspired by the work of Jaiswal et al.,^1^ which shows that a subset of bone marrow-derived microglial cells, the myeloid-monocytic cells of the central nervous system, can carry CH-associated somatic mutations, we interrogated our *MutDetect-Seq* single cell RNA sequencing profiles in non-AD patients with the most common CH-driver mutation, DNMT3A. Since microglia play a crucial role in the maintenance of brain homeostasis by removal of cellular debris and un- or misfolded proteins via phagocytosis^5^ and microglia phagocytosis decreases during aging, we hypothesized that an augmented phagocytotic capacity of mutant cells might contribute to the ameliorative effects on neurodegenerative processes. Indeed, as shown in the figure, monocytes harbouring a DNMT3A CH mutation displayed a pronounced pro-phagocytic phenotype at single-cell transcriptional level compared to wild-type cells, illustrated by a significant upregulation of gene ontology pathways associated with phagocytosis in general as well as microglia-specific (fig. 1b). Specifically, some prominent examples of upregulated pro-phagocytotic genes, like calreticulin (CALR), which exerts important functions in protein folding control, cytochrome B alpha chain (CYBA), the primary component of the microbicidal oxidase system of phagocytosis, Fc receptors important for proteasome-mediated degradation (FCER1G) and phagocytic cup formation for target degradation (FCGR1A), as well as the calcium- and zinc-binding proteins S100A8/A9, which exert a plethora of immune surveillance functions including their action as danger associated molecular pattern (DAMP) molecules, were increased in DNMT3A mutant cells and the regulation was further validated in vitro by siRNA mediated silencing DNMT3A in monocyte-derived macrophages (fig 1d). Thus, if human bone marrow-derived monocytic cells indeed invade the brain and adopt a microglia-like phenotype, cells carrying CH DNMT3A mutations may replenish the brain with increased phagocytotic capacity counteracting the age-dependent decline of microglia phagocytosis, thereby playing a causative role in reducing neurodegenerative processes by phagocytosing ß-amyloid plaques and neurofibrillary tangles - the hallmark neuropathological features in AD.^3^

**Fig. 1.**
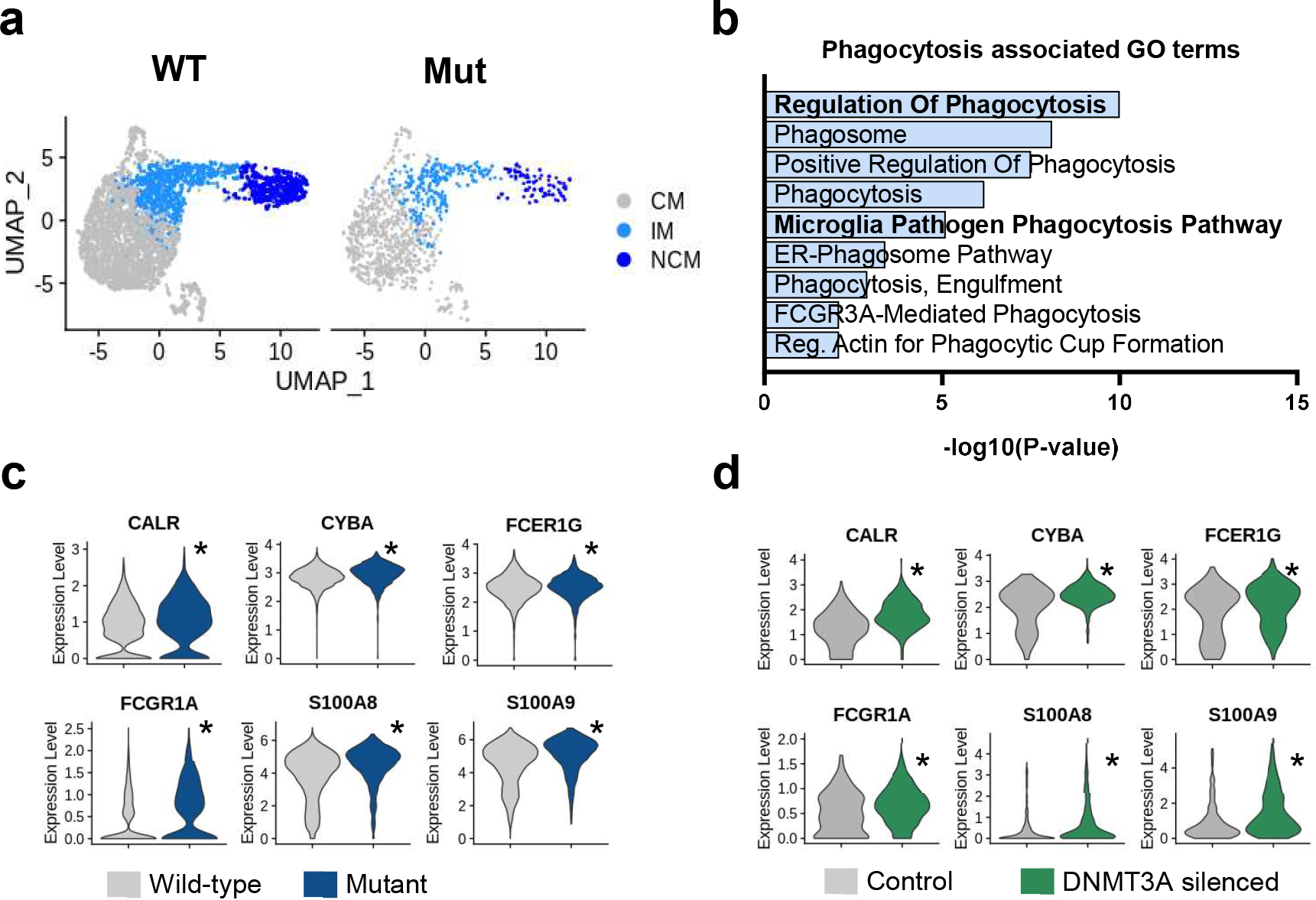
DNMT3A mutant monocytes show a pronounced pro-phagocytosis phenotype. a, Identification and distribution of monocyte subclasses with and without mutations (7,041 monocytes (total): 3,614 WT CMs and 821 mutant CMs; 1,328 WT IMs and 272 mutant IMs; and 906 WT NCMs and 100 mutant NCMs). d, Upregulated genes in DNMT3A mutant monocytes relative to wild-type call gene ontology terms associated with phagocytosis. c/d, Relative expression of upregulated genes in (c) circulating mutant monocytes of HF patients versus wild type cells and (d) in vitro validation of phagocytosis upregulated genes in DNMT3A silenced macrophages. (n = 644 control cells, n = 387 DNMT3A silenced cells). (Significance denoted by asterisk (*) and determined for c,d by Wilcoxon Rank Sum test, with adjusted p-value <0.05).

In summary, deciphering the phenotypic differences between mutant and wild-type monocytes disclosed by a novel method for assessing cell-intrinsic effects of CH may not only provide important insights into a potential mechanism underlying the unexpected role of CH in slowing the progression of AD, but may also provide valuable information for understanding the effects of different CH mutations in a disease-specific context.

## Data Availability

All data produced are available online at https://www.ebi.ac.uk/biostudies/arrayexpress/studies/E-MTAB-13016

